# Association Between Serum CtBP2 Levels and Obesity Markers: A Cross-Sectional Analysis of Metabolic Syndrome Components

**DOI:** 10.64898/2026.05.16.26353386

**Authors:** Oumo David, Namasinga Agatha, Amoding Martha Ikwap, Ekalu Moses, Mpumwire Peninah

## Abstract

**Background:** C-terminal binding protein 2 (CtBP2) has been implicated in metabolic regulation, but its association with specific measures of adiposity and lipid profiles in humans remains unclear. This study examined the relationship between circulating CtBP2 levels and key components of metabolic syndrome, focusing on body fat distribution and lipid markers.

**Methods:** Data from 508 participants (259 men, 249 women) from a publicly available dataset were analyzed. Serum CtBP2 concentrations were measured using ELISA. Associations with obesity markers (BMI, waist circumference, waist-to-hip ratio) and lipid profiles (triglycerides, HDL cholesterol) were assessed using Spearman correlation and linear regression, adjusting for age and sex.

**Results:** CtBP2 levels showed weak but statistically significant positive correlations with all measures of adiposity, with the strongest association observed for waist circumference (ρ = 0.150, p < 0.001), followed by BMI (ρ = 0.120, p = 0.007) and waist-to-hip ratio (ρ = 0.098, p = 0.027). No significant correlations were found with triglycerides or HDL cholesterol. In the regression model predicting BMI, age, and sex were significant predictors, while CtBP2 demonstrated a trend toward association (β = 0.080, p = 0.052).

**Conclusion:** Circulating CtBP2 appears to be modestly associated with measures of adiposity, particularly abdominal fat, but not with lipid abnormalities. These findings suggest a potential role for CtBP2 in obesity-related metabolic dysregulation and underscore the need for further mechanistic studies to clarify its clinical relevance.

## Background

Metabolic syndrome (MetS) refers to a cluster of metabolic abnormalities that occur together and significantly increase the risk of cardiovascular disease and type 2 diabetes. These abnormalities commonly include central obesity, insulin resistance, hypertension, and dyslipidemia. The global prevalence of MetS has increased considerably over the past few decades, largely driven by rising obesity rates, sedentary lifestyles, and dietary changes [1, 2]. As the burden of metabolic disorders continues to grow, there is growing interest in identifying the molecular mechanisms underlying the development and progression of MetS.

Among the molecular regulators that have recently attracted attention is C-terminal binding protein 2 (CtBP2), a transcriptional co-repressor that regulates gene expression through interactions with multiple transcription factors and chromatin-modifying complexes. CtBP2 was initially studied extensively in cancer biology because of its ability to repress tumor suppressor genes and influence cell proliferation [3]. However, emerging evidence suggests that CtBP2 also participates in several essential biological processes beyond tumorigenesis, including cellular differentiation, apoptosis, and metabolic regulation [4]. Despite these insights, the precise role of CtBP2 in metabolic disorders remains incompletely understood.

Several studies suggest that CtBP2 may influence metabolic pathways relevant to the development of metabolic syndrome. In particular, CtBP2 has been implicated in adipocyte differentiation and lipid metabolism. It interacts with key transcriptional regulators such as peroxisome proliferator-activated receptor gamma (PPARγ), which plays a central role in adipogenesis and fat storage [5, 6]. Dysregulation of these pathways may contribute to abnormal adipose tissue accumulation and central obesity, both of which are defining features of MetS. In addition, CtBP2 has been shown to interact with sterol regulatory element-binding proteins, transcription factors that regulate genes involved in cholesterol and triglyceride synthesis. Since dyslipidemia, characterized by elevated triglycerides and reduced high-density lipoprotein cholesterol, is a major component of MetS, these interactions suggest that CtBP2 may play an important role in lipid homeostasis [3, 7].

Beyond its potential effects on adipose tissue and lipid metabolism, CtBP2 may also contribute to the inflammatory and oxidative processes associated with metabolic syndrome. Chronic low-grade inflammation is widely recognized as a major mechanism underlying insulin resistance and cardiometabolic complications [8, 9]. Experimental studies have indicated that CtBP2 can influence the expression of several pro-inflammatory cytokines, including interleukin-6 (IL-6) and tumor necrosis factor-alpha (TNF-α). These cytokines are commonly elevated in individuals with MetS and are believed to contribute to metabolic dysregulation and disease progression [10-12]. Therefore, CtBP2 may represent an important molecular link between obesity-related metabolic disturbances and inflammatory signaling pathways.

Despite these emerging findings, limited research has examined the relationship between CtBP2 and the clinical features of metabolic syndrome in human populations. A clearer understanding of this relationship may provide insights into the molecular mechanisms underlying metabolic dysfunction and help identify potential biomarkers or therapeutic targets. Therefore, the present study aimed to investigate the association between circulating CtBP2 levels and key components of metabolic syndrome, including obesity indicators such as body mass index, waist circumference, and waist-to-hip ratio, as well as lipid profile abnormalities such as elevated triglycerides and reduced HDL cholesterol. Understanding these associations may help clarify the potential role of CtBP2 in the development of metabolic syndrome.

## Methods

### Data Source and Study Population

This study used a publicly available dataset obtained from Mendeley Data, originally published by Gaonian Zhao in 2025 [13]. The dataset was generated to investigate the relationship between circulating levels of C-terminal binding protein 2 and type 2 diabetes mellitus (T2DM).

The dataset included 508 participants, comprising 256 individuals with newly diagnosed T2DM and 252 age- and sex-matched controls with normal glucose tolerance. Serum CtBP2 concentrations were measured for all participants using an enzyme-linked immunosorbent assay (ELISA). In addition to CtBP2 measurements, the dataset contained demographic, anthropometric, and biochemical variables relevant to metabolic health.

### Variables

For the present analysis, the primary exposure variable was serum CtBP2 concentration. Anthropometric indicators of obesity included body mass index (BMI), waist circumference (WC), and waist-to-hip ratio (WHR). Lipid profile indicators included triglycerides (TG) and high-density lipoprotein cholesterol (HDL-C). Age and sex were included as covariates.

### Statistical Analysis

All statistical analyses were conducted using R (version 4.4.2). Descriptive statistics were calculated for continuous variables and summarized using means, medians, and standard deviations where appropriate.

Because several variables did not meet the assumptions of normal distribution, Spearman’s rank correlation coefficient was used to evaluate associations between serum CtBP2 levels and metabolic parameters, including BMI, WC, WHR, TG, and HDL-C.

To further examine the relationship between CtBP2 and obesity, a linear regression model was fitted with BMI as the dependent variable and serum CtBP2 level as the main predictor, adjusting for age and sex. Model assumptions, including linearity, homoscedasticity, and normality of residuals, were assessed using standard diagnostic plots.

Data manipulation and visualization were performed using the R packages dplyr and ggplot2, while statistical modeling and result reporting used base R functions and the broom package.

### Ethical Considerations

The original study received ethical approval from its Institutional Review Board, and all participants provided written informed consent prior to participation. The dataset used in the present study was fully anonymized and publicly available. Therefore, additional ethical approval was not required for this secondary data analysis.

## Results

### Study Population Characteristics

The study included 508 participants, comprising 259 men and 249 women. Serum CtBP2 concentrations showed substantial variability, with a mean level of 3.44 U/L (range: 0.22–25.34 U/L).

The mean body mass index (BMI) was 24.76 kg/m^2^, while the mean waist circumference (WC) was 91.04 cm, indicating moderate adiposity within the cohort. The average triglyceride (TG) concentration was 1.94 mmol/L, and the mean high-density lipoprotein cholesterol (HDL-C) level was 1.21 mmol/L. The characteristics of the study population are shown in Table 1.

**Table 1.** Descriptive statistics for the continuous variables.

| Variable | Min | 1<sup>st</sup> Qu | Median | Mean | 3<sup>rd</sup> Qu | Max |
| --- | --- | --- | --- | --- | --- | --- |
| BMI | 18.55 | 22.26 | 24.30 | 24.76 | 26.91 | 34.82 |
| CtBP2 | 0.22 | 1.27 | 2.37 | 3.44 | 3.96 | 25.34 |
| HDL | 0.53 | 1.01 | 1.20 | 1.21 | 1.39 | 2.16 |
| TG | 0.36 | 0.99 | 1.40 | 1.94 | 2.29 | 14.05 |
| WC | 71.30 | 84.30 | 89.15 | 91.04 | 96.33 | 121.0 |
| WHR | 0.78 | 0.87 | 0.92 | 0.92 | 0.96 | 1.06 |

### Association Between CtBP2 and Obesity Markers

Spearman correlation analysis demonstrated weak but statistically significant positive associations between serum CtBP2 levels and measures of adiposity. The strongest association was observed between CtBP2 and waist circumference (ρ = 0.150, p < 0.001). CtBP2 also showed a positive correlation with BMI (ρ = 0.120, p = 0.007) and waist-to-hip ratio (WHR) (ρ = 0.098, p = 0.027). These results indicate that higher CtBP2 levels were modestly associated with greater overall and central adiposity. Scatterplots illustrating these relationships are shown in Figure 1.

**Figure 1.**
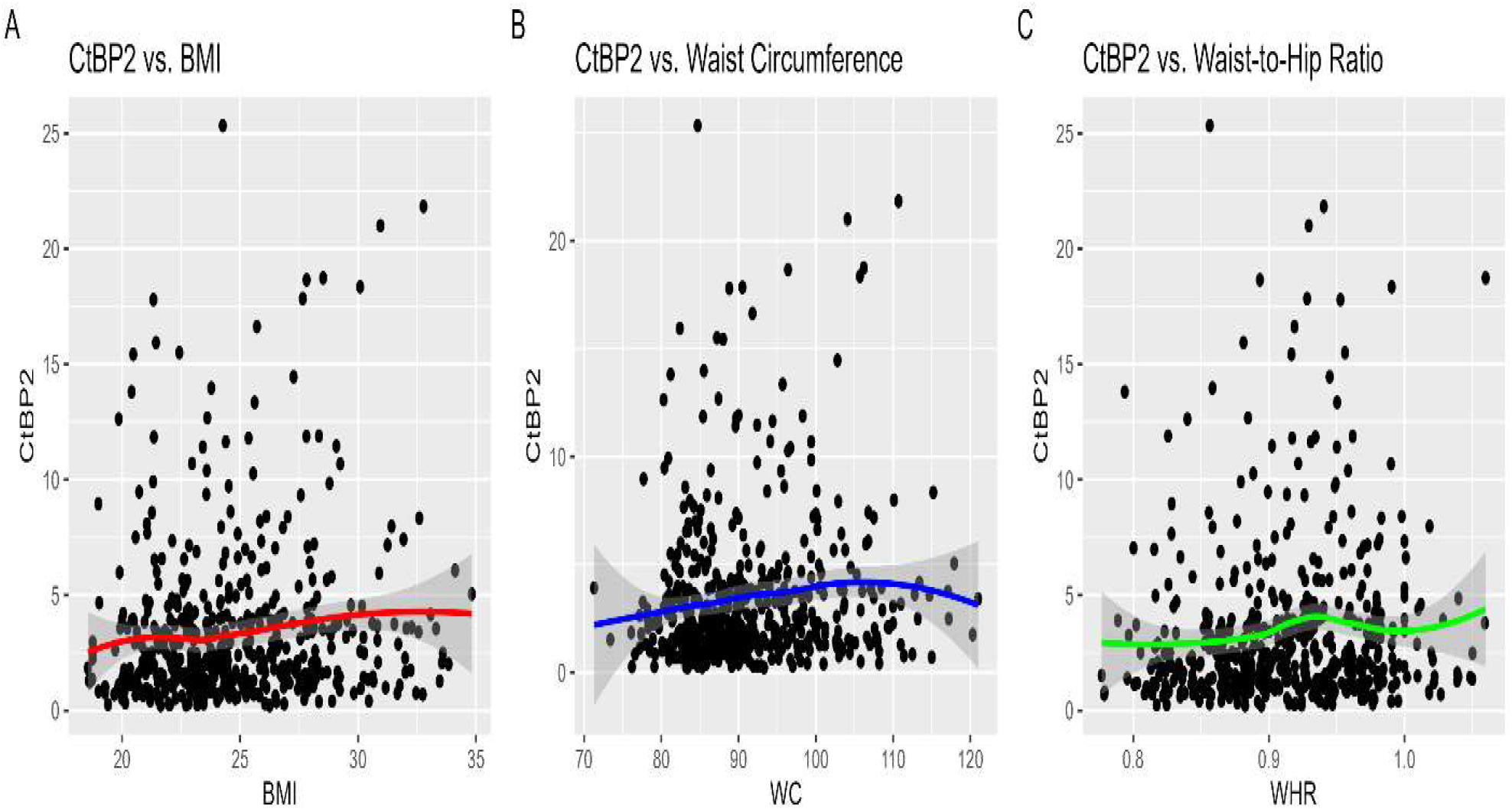
Scatterplots illustrating the relationships between CtBP2 and obesity markers. (A) BMI, (B) Waist Circumference (WC), and (C) Waist-to-Hip Ratio (WHR). Loess smoother lines indicate the trend, and Spearman’s correlation coefficients (ρ)

### Association Between CtBP2 and Lipid Profile Markers

No statistically significant associations were observed between CtBP2 levels and lipid profile indicators. The correlation between CtBP2 and triglycerides (TG) was weak and not statistically significant (ρ = 0.083, p = 0.062). Similarly, the correlation between CtBP2 and HDL-C was minimal and not significant (ρ = −0.035, p = 0.437). These findings suggest that CtBP2 levels were not strongly associated with lipid abnormalities in this cohort. The corresponding scatterplots are presented in Figure 2.

**Figure 2.**
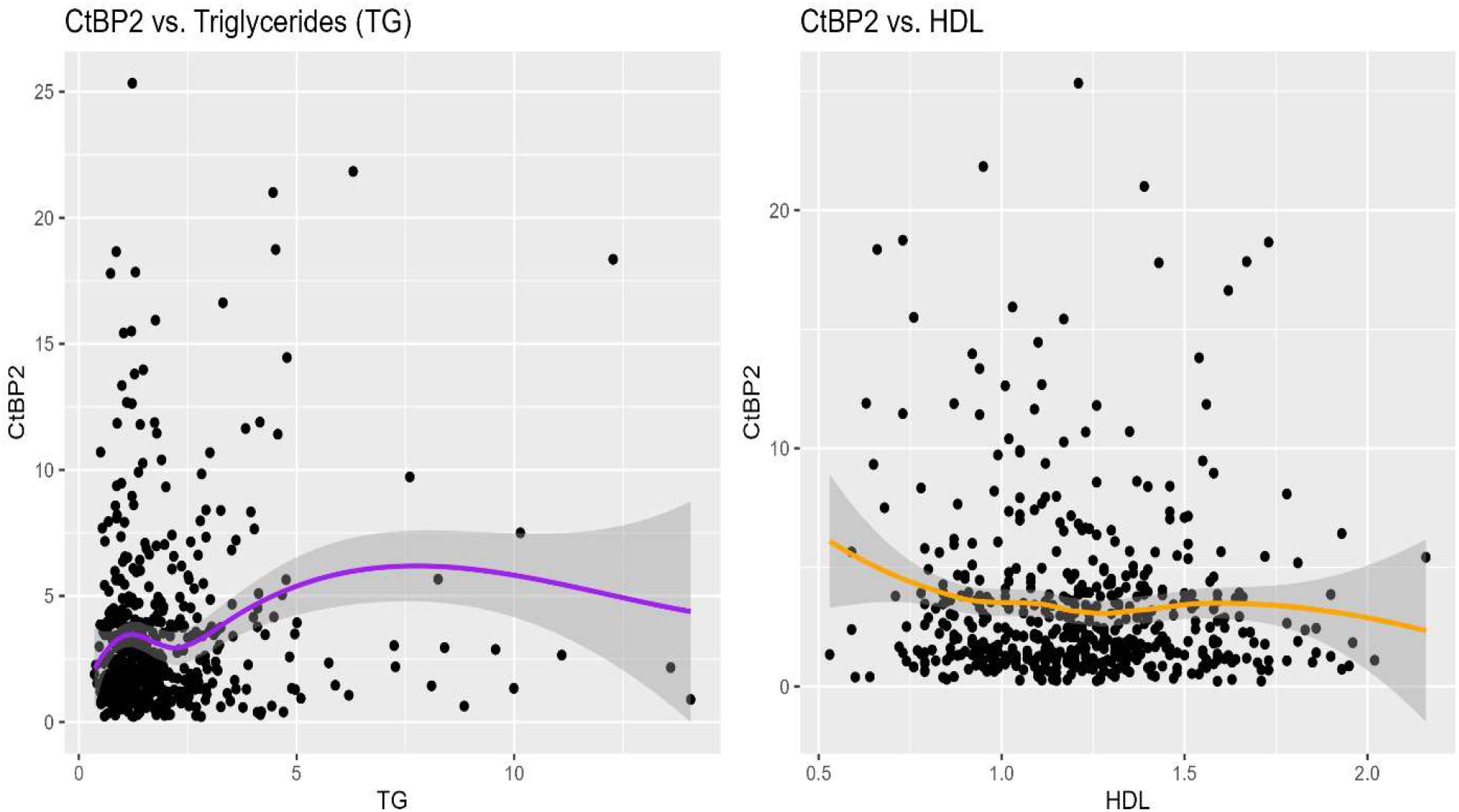
Scatterplots showing the relationship between CtBP2 and lipid profile markers. (A) CtBP2 vs. TG, (B) CtBP2 vs. HDL. Loess smoothers (purple and orange lines) indicate the trend of the relationships. Spearman’s correlation coefficients (ρ) and p-values are shown for each plot

#### Linear Regression Analysis

A multiple linear regression model was fitted to examine the association between CtBP2 and BMI, adjusting for age and sex. The model explained a small proportion of the variance in BMI (R^2^ = 0.057, p < 0.001). As shown in Table 2, age was negatively associated with BMI (β = −0.039, p = 0.002), indicating that older participants tended to have lower BMI values. Sex was also a significant predictor, with females exhibiting lower BMI values compared to males (β = −1.007, p < 0.001). After adjustment for these variables, CtBP2 showed a positive association with BMI, although this relationship did not reach conventional statistical significance (β = 0.080, p = 0.052).

**Table 2.** Linear Regression Model Predicting BMI.

| Predictor | Estimate ($\beta$) | Std_Error | t_value | p_value |
| --- | --- | --- | --- | --- |
| (Intercept) | 27.990 | 0.770 | 36.35 | <0.001 |
| CtBP2 | 0.080 | 0.041 | 1.95 | 0.052 |
| Age | -0.039 | 0.012 | -3.13 | 0.002 |
| Sex | -1.007 | 0.293 | -3.43 | <0.001 |

## Discussion

This study investigated the relationship between circulating CtBP2 levels and key components of metabolic syndrome, with particular focus on markers of adiposity and lipid metabolism. The findings suggest that CtBP2 is modestly associated with indicators of obesity, especially measures of central adiposity, while showing little relationship with lipid profile markers.

### CtBP2 And Obesity Markers

The analysis revealed weak but statistically significant positive correlations between serum CtBP2 levels and several measures of adiposity, including BMI and waist circumference. Among these, the association with waist circumference was the strongest, suggesting a closer relationship between CtBP2 and central fat accumulation. Central obesity is a well-recognized driver of metabolic syndrome and cardiometabolic disease, making this observation biologically relevant.

These findings are consistent with experimental evidence suggesting that CtBP2 functions as a transcriptional co-repressor involved in metabolic regulation, particularly in pathways related to adipocyte differentiation and energy balance. Previous studies have shown that CtBP2 interacts with transcription factors that regulate adipogenesis, including those involved in PPARγ signaling and other regulators of fat storage. Through these mechanisms, CtBP2 may influence adipose tissue development and metabolic homeostasis [3, 6, 9].

The regression analysis further suggested a positive association between CtBP2 and BMI after adjusting for age and sex, although the relationship narrowly missed conventional statistical significance. While the effect size was small, the direction of the association is consistent with the observed correlations and may indicate that CtBP2 contributes modestly to variations in adiposity. The stronger association with waist circumference than with BMI may indicate that CtBP2 is more closely linked to visceral fat accumulation than to overall body mass [14, 15].

### CtBP2 And Lipid Profile

In contrast to the findings for adiposity markers, CtBP2 levels were not significantly associated with lipid profile indicators in this cohort. Neither triglyceride levels nor HDL cholesterol showed meaningful correlations with CtBP2. This suggests that CtBP2 may not directly influence circulating lipid levels, at least within the context of the present dataset.

These results differ somewhat from experimental studies that have proposed a role for CtBP2 in lipid metabolism, particularly through interactions with sterol regulatory element-binding proteins (SREBPs), which regulate genes involved in cholesterol and triglyceride synthesis. The absence of significant associations in the present analysis may reflect differences between experimental models and human populations, or it may indicate that CtBP2 influences lipid metabolism indirectly rather than through measurable changes in circulating lipid levels [16, 17].

### Predictors of BMI

The regression model identified age and sex as significant predictors of BMI, with older individuals and females demonstrating lower BMI values on average. These patterns are consistent with demographic trends reported in epidemiological studies, where obesity prevalence often varies by age and sex due to hormonal, behavioral, and lifestyle factors.

Although CtBP2 was included in the model as a potential predictor of BMI, its contribution was relatively small. The modest explanatory power of the model (R^2^ = 0.057) indicates that BMI is influenced by a wide range of biological and environmental factors that were not captured in this analysis. Nevertheless, the observed trend suggests that CtBP2 may still play a minor role in metabolic regulation, warranting further investigation [18, 19].

## Conclusion

This study explored the association between circulating CtBP2 levels and key components of metabolic syndrome. The findings indicate that CtBP2 is modestly associated with markers of adiposity, particularly waist circumference, suggesting a potential relationship with abdominal obesity. In contrast, no significant associations were observed between CtBP2 and lipid profile indicators, including triglycerides and HDL cholesterol.

These results suggest that CtBP2 may play a limited but potentially relevant role in pathways related to adiposity rather than lipid metabolism. However, the observed associations were relatively weak, indicating that CtBP2 is unlikely to be a major determinant of metabolic syndrome components on its own.

Further research, particularly longitudinal and mechanistic studies, is needed to clarify the biological pathways linking CtBP2 to metabolic regulation and to determine whether CtBP2 could serve as a biomarker or therapeutic target in metabolic disorders.

## Limitations

The low R^2^ values in the regression models suggest that unmeasured factors, such as diet, physical activity, and genetic predisposition, may influence the variability in BMI and other outcomes. The study population may not reflect broader populations, limiting the extent to which the findings can be applied.

## Future Directions

Future research should investigate the mechanistic pathways linking CtBP2 to obesity and metabolic dysregulation. For example, experimental studies could examine whether CtBP2 influences adipogenesis, lipid metabolism, or insulin signaling. Additionally, longitudinal studies are necessary to determine the temporal relationship between CtBP2 levels and components of metabolic syndrome. Subgroup analyses by gender, age, or metabolic health status could also offer further insights into the role of CtBP2 in different populations.

## Declarations

### Data availability

The original dataset is publicly available on Mendeley Data: https://data.mendeley.com/datasets/x469xfg5j8/1

All analysis code and derived data are available upon request from the authors.

### Author Contributions

OD and MP conceived the research idea. AMI, EK, and MP contributed to refining the study design and methods. OD and NA were actively involved in data analysis and interpretation. NA, AMI, and EK drafted the manuscript. OD, NA, and EK were involved in the manuscript review. All the authors reviewed and approved the final manuscript.

### Consent for publication

Not applicable

### Ethical Approval

Not applicable (secondary analysis of publicly available data).

## Acknowledgements

The authors gratefully acknowledge Gaonian Zhao for making the dataset publicly available on Mendeley Data, which enabled this secondary analysis. We also extend our appreciation to the original researchers and the participants whose data underpin this work.

## References

1. Swarup S AI, Grigorova Y, et al. : Metabolic Syndrome. StatPearls [Internet] 2024.

2. Saklayen MG: The Global Epidemic of the Metabolic Syndrome. Curr Hypertens Rep 2018, 20(2):12.

3. Sekiya M, Kainoh K, Sugasawa T, Yoshino R, Hirokawa T, Tokiwa H, Nakano S, Nagatoishi S, Tsumoto K, Takeuchi Y et al: The transcriptional corepressor CtBP2 serves as a metabolite sensor orchestrating hepatic glucose and lipid homeostasis. Nature Communications 2021, 12(1):6315.

4. Blevins MA, Huang M, Zhao R: The Role of CtBP1 in Oncogenic Processes and Its Potential as a Therapeutic Target. Mol Cancer Ther 2017, 16(6):981–990.

5. Hernandez-Quiles M, Broekema MF, Kalkhoven E: PPARgamma in Metabolism, Immunity, and Cancer: Unified and Diverse Mechanisms of Action. Frontiers in Endocrinology 2021, 12.

6. Saito K, Sekiya M, Kainoh K, Yoshino R, Hayashi A, Han S-I, Araki M, Ohno H, Takeuchi Y, Tsuyuzaki T et al: Obesity-induced metabolic imbalance allosterically modulates CtBP2 to inhibit PPAR-alpha transcriptional activity. Journal of Biological Chemistry 2023, 299(7):104890.

7. Chandrasekaran P, & Weiskirchen, R. : The Role of SCAP/SREBP as Central Regulators of Lipid Metabolism in Hepatic Steatosis. . International Journal of Molecular Sciences, 25(2), 1109 2024.

8. Wang W, Tu M, Qiu XP, Tong Y, Guo XL: The Interplay of Systemic Inflammation and Oxidative Stress in Connecting Perirenal Adipose Tissue to Hyperuricemia in Type 2 Diabetes Mellitus: A Mediation Analysis. J Inflamm Res 2024, 17:11319–11329.

9. Sekiya M, Kainoh K, Saito K, Yamazaki D, Tsuyuzaki T, Chen W, Kobari Y, Nakata A, Babe H, Shimano H: C-Terminal Binding Protein 2 Emerges as a Critical Player Linking Metabolic Imbalance to the Pathogenesis of Obesity. J Atheroscler Thromb 2024, 31(2):109–116.

10. Bashir H, Ahmad Bhat S, Majid S, Hamid R, Koul RK, Rehman MU, Din I, Ahmad Bhat J, Qadir J, Masood A: Role of inflammatory mediators (TNF-α, IL-6, CRP), biochemical and hematological parameters in type 2 diabetes mellitus patients of Kashmir, India. Med J Islam Repub Iran 2020, 34:5.

11. Li H, Zhang C, Bian L, Deng H, Blevins M, Han G, Fan B, Yang C, Zhao R, High W et al: Inhibition of CtBP-Regulated Proinflammatory Gene Transcription Attenuates Psoriatic Skin Inflammation. J Invest Dermatol 2022, 142(2):390–401.

12. Chen Z, Dong WH, Qiu ZM, Li QG: The Monocyte-Derived Exosomal CLMAT3 Activates the CtBP2-p300-NF-κB Transcriptional Complex to Induce Proinflammatory Cytokines in ALI. Mol Ther Nucleic Acids 2020, 21:1100–1110.

13. Zhao G: A dataset pertaining to the investigation of serum CtBP2 levels in relation to Type 2 Diabetes Mellitus. Mendeley Data 2025, 1.

14. Paley CA, Johnson MI: Abdominal obesity and metabolic syndrome: exercise as medicine? BMC Sports Science, Medicine and Rehabilitation 2018, 10(1):7.

15. Barroso TA, Marins LB, Alves R, Gonçalves ACS, Barroso SG, Rocha GdS: Association of Central Obesity with The Incidence of Cardiovascular Diseases and Risk Factors. International Journal of Cardiovascular Sciences 2017, 30(5):416–424.

16. Sekiya M, Ma Y, Kainoh K, Saito K, Yamazaki D, Tsuyuzaki T, Chen W, Adi Putri PIP, Ohno H, Miyamoto T et al: Loss of CtBP2 may be a mechanistic link between metabolic derangements and progressive impairment of pancreatic β cell function. Cell Reports 2023, 42(8):112914.

17. Ferrarese R, Izzo A, Andrieux G, Lagies S, Bartmuss JP, Masilamani AP, Wasilenko A, Osti D, Faletti S, Schulzki R et al: ZBTB18 inhibits SREBP-dependent lipid synthesis by halting CTBPs and LSD1 activity in glioblastoma. Life Sci Alliance 2023, 6(1).

18. Aslani A, Faraji A, Allahverdizadeh B, Fathnezhad-Kazemi A: Prevalence of obesity and association between body mass index and different aspects of lifestyle in medical sciences students: A cross-sectional study. Nurs Open 2021, 8(1):372–379.

19. Ng M, Dai X, Cogen RM, Abdelmasseh M, Abdollahi A, Abdullahi A, Aboagye RG, Abukhadijah HJ, Adeyeoluwa TE, Afolabi AA et al: National-level and state-level prevalence of overweight and obesity among children, adolescents, and adults in the USA, 1990–2021, and forecasts up to 2050. The Lancet 2024, 404(10469):2278–2298.

